# Epidemiology of RSV-A and RSV-B in Adults and Children with Medically-Attended Acute Respiratory Illness over Three Seasons

**DOI:** 10.1101/2022.11.04.22281968

**Authors:** Katherine M. Begley, Aleda M. Leis, Joshua G. Petrie, Rachel Truscon, Emileigh Johnson, Erin McSpadden, Lois E Lamerato, Melissa Wei, Arnold S. Monto, Emily T. Martin

**Author notes:** **Corresponding Author:** Emily T. Martin, PhD MPH, University of Michigan School of Public Health, 1415 Washington Heights, Ann Arbor, Michigan 48109. Center for Clinical Epidemiology & Population Health, Marshfield Clinic Research Institute, Marshfield, WI.

## Abstract

**Background:** RSV is a frequent cause of respiratory illness less often diagnosed outside hospital settings; thus, overall prevalence of RSV-associated illness is under-recognized. Information about presence of RSV among those with chronic conditions is especially needed with recent advances in vaccine development.

**Methods:** Participants prospectively enrolled in an ambulatory surveillance study of respiratory illness (MFIVE) were tested by RT-PCR for RSV and influenza. Participant and illness characteristics were collected by in-person survey and EMR review. Chronic conditions were characterized by the Multimorbidity-weighted index (MWI). Viral factors, including subtype and viral load, were compared between RSV-A and RSV-B. Multivariate logistic regression models were used to compare participant and illness characteristics between those with RSV and those with influenza. Comparisons were also made across RSV subtypes.

**Results:** Among 4,442 individuals enrolled in MFIVE from fall 2017 to spring 2020, 9.9% (n=441) had RSV detected. RSV+ participants with increased viral load had increased odds of illness lasting ≥ 7 days [OR_adj_=2.39 (95% CI: 1.03-5.51) p-value=0.04]. Adults with RSV had higher median MWI scores compared to influenza and RSV/influenza-negative (1.62, 0.40, 0.64, respectively).

**Conclusions:** Our findings support the need for ongoing RSV surveillance, particularly in older adults and those with multimorbidity. Our findings support a recognition of multimorbidity as a significant contributor to RSV-associated MAARI among outpatient adults, with particularly notable impacts among adults under 65.

## Introduction

Respiratory Syncytial Virus (RSV) is one of the leading global causes of acute respiratory illness (ARI) among children and is known to be a significant cause of ARI among high-risk adults, particularly those with underlying cardiopulmonary conditions [1–5]. Assessments of multi-year patterns of RSV-A and RSV-B epidemiology in both children and adults with medically attended acute respiratory illness (MAARI) are needed for the outpatient setting to inform the need for and use of future RSV vaccines.

Older adults experience a higher incidence of RSV-associated MAARI as well as severe RSV-associated illness outcomes, including hospitalization, when compared to younger adults [6–9]. An individual’s health history – such as age and underlying chronic conditions, especially among adults – and characteristics of the virus – including subtype and viral load – may contribute to increased RSV severity. The prevalence of multimorbidity – the coexistence of multiple underlying chronic conditions – increases significantly with age. As individuals with multimorbidity are more likely to experience adverse health outcomes [10–14], multimorbidity may also increase the risk of respiratory virus infection or the outcomes of medically-attended acute respiratory illness.

The primary objective of this study was to describe characteristics and illness outcomes among adults and children with MAARI at outpatient clinics located in southeast Michigan between 2017 and 2020. To do so, we evaluated the relationship between quantitative viral load and illness outcomes and whether viral subtype modified these associations and described the impact of multimorbidity on RSV illness in adults.

## Methods

### Source Population

Data and specimens for this study were available from three respiratory virus seasons–2017/18, 2018/19, and 2019/20–of the Michigan Henry Ford Influenza Vaccine Effectiveness (MFIVE) study. Data collection was ended on March 13, 2020 due to COVID-19 mitigation. MFIVE is an ongoing, prospective, ambulatory-care study in southeast Michigan with coverage across 19 (2017/18), 23 (2018/19), and 22 (2019/20) outpatient clinics. MFIVE enrolls over 1,000 people annually with MAARI for analysis of influenza vaccine effectiveness [15–18]. Patients six months of age and older presenting with medically attended acute respiratory illness (MAARI) meeting a standard case definition (lasting ≤ seven days with a cough) and who have not taken influenza antiviral treatment for their current illness were eligible for participation.

### Data collection

Following informed consent, MFIVE research staff conducted an enrollment interview to collect demographic and household characteristic data, information on their current illness and overall health status, and vaccination status. Staff collected throat and nasal swab specimens and delivered them to the Michigan Center for Respiratory Virus Research and Response for processing and storage. Seven days post-enrollment, participants received an online follow-up questionnaire to complete at home. Through this survey, participants self-reported illness duration and recovery metrics, including subsequent care-seeking behavior. The University of Michigan and Henry Ford Health Institutional Review Boards provided ethical approval for this work.

### Illness Characteristics

The number of symptoms reported–fever, sore throat, congestion–were operationalized dichotomously for assessing symptom burden (low 0-1 symptoms vs. high 2-3 symptoms), using any combination of symptoms reported. Length of illness was determined using self-reported illness onset date recorded at enrollment and recovery date from the follow-up survey and then dichotomized as extended illness (≥ 7 days) or not (< 7 days). Variables for subsequent seeking of medical care or treatment were combined and dichotomized (sought subsequent treatment, yes/no) and included visiting a doctor’s office, urgent care clinic, retail pharmacy clinic, or emergency department/hospital.

### Clinical Data

Electronic Health Record (EHR) data were abstracted to obtain ICD-10 diagnosis codes indicating high-risk conditions up to one year prior to enrollment, and all ICD-10 codes recorded by the healthcare provider seen at enrollment. Participant date of birth, sex and height (centimeters) and weight (kilograms) closest to enrollment for calculating BMI (kg/m^2^) were also abstracted.

### Viral Characteristics

Viral RNA was extracted with QiaAmp Viral RNA mini kits (Qiagen, Germany). RSV, influenza, and other viral respiratory pathogens (Rhinovirus, Parainfluenza, Human metapneumovirus, Seasonal coronaviruses, Bocavirus, Adenovirus, and Enterovirus) were detected by RT-PCR using Fast Track Diagnostics real-time multiplex PCR respiratory panel (Siemens Healthineer Company, Luxembourg). Sample viral subtype and quantitative viral load were determined by Real-Time PCR (RT-PCR) on an ABI 7500 instrument (Thermo Fisher Scientific), protocol described elsewhere [19,20]. All test plates included nuclease-free water as a negative control as well as six RNA transcript standards, ranging from 1 × 10^4^ – 1 × 10^9^ copies of viral RNA. Viral load was determined through comparing unknown samples to transcript standards using curves generated by qPCR.

### Multimorbidity-weighted index (MWI)

To determine adult multimorbidity status, the main exposure, we applied the Multimorbidity Weighted Index (MWI), a validated, patient-centric measure of multimorbidity adapted for ICD-10 compatibility [21,22]. The MWI measures the impact of underlying conditions on patient physical functioning. Ninety-five conditions were represented in the MWI across the following categories: cardiovascular, endocrine, gastrointestinal, hematologic, immunologic, integumentary, musculoskeletal, nervous, oncologic, ophthalmologic, oral, psychiatric, pulmonary, renal, and reproductive [21].

All ICD-10 diagnosis codes extracted from participant EHR were included to construct individual MWI scores using available macros on SAS software version 9.4 (SAS Institute, Cary NC). For the primary analysis we evaluated multimorbidity continuously and dichotomously (>0 versus 0 MWI), where indicated.

### Statistical Analysis

Participants with RSV were compared to two groups: participants positive for influenza and participants negative for both RSV and influenza. Overall descriptive statistics were calculated for all eligible study participants.

For statistical analyses, data from all seasons were pooled to increase power, and study year was included in regression models for pooled data. Age was categorized using epidemiologically meaningful cut points: 0-4 years, 5-17 years, 18-49 years, 50-64 years, and ≥ 65 years, respectively). Race (White, Black, and Other), education status (Less than High School, Graduated High School/GED, Some College, Bachelor’s Degree, and Advanced Degree) and obesity (BMI ≥ 30). Time between illness onset and specimen collection was categorized (0-2 days and 3+ days). Viral load was log10-transformed for analysis as a continuous variable as well as dichotomized using median viral load as the cut point. Using median sample viral load as a threshold, samples with a quantitative viral load ≥ 2.2×10^4^ copies/mL (log10-transformed ≥ 4.3) were compared to samples with a quantitative viral load < 2.2×10^4^ copies/mL (log10-transformed < 4.3).

For viral characteristic analyses, all RSV-positive participants with subtype and viral load data available were included. Pearson’s *r* correlation coefficient was used to assess the association between log viral load and viral subtype. The Mann-Whitney U Test was used to detect differences in viral load (copies/mL) between RSV-A and RSV-B samples.

To test the association between quantitated viral load and RSV-associated illness outcomes, both univariate and multivariable Firth logistic regression models were constructed, and odds ratios reported. Multivariable models were adjusted for age, sex, race, season, and time between illness onset and specimen collection. Firth-adjusted regression models were used to account for potential bias from smaller outcome counts in stratified analyses [23]. We provide overall and age-stratified estimates, where indicated, as well as estimates stratified by viral subtype to assess effect modification. To test for effect modification more precisely, a statistical interaction term between viral load and viral subtype was included in all overall illness outcome models.

For multimorbidity analyses, only adults ICD codes were included. Participants under the age of 18 were excluded from multimorbidity analyses as the MWI is not validated for use in children. Descriptive statistics were calculated separately for adults included in multimorbidity analyses. Firth-adjusted multivariate logistic regression models were used to assess the association between multimorbidity and ARI outcomes, adjusting for age, sex, race, and season. All statistical analyses were conducted on SAS software version 9.4 (SAS Institute, Cary NC).

## Results

### Epidemiology

From 2017-2020, 4,490 people were enrolled in MFIVE; 16 participants were excluded from this analysis due to inconclusive RSV testing and 32 participants were excluded due to RSV-influenza co-detection (Figure 1). Overall (n=4,442), 441 study-eligible cases of RSV, 1,341 cases of influenza, and 2,660 participants negative for both RSV and influenza were included (Figure 2).

**Figure 1.**
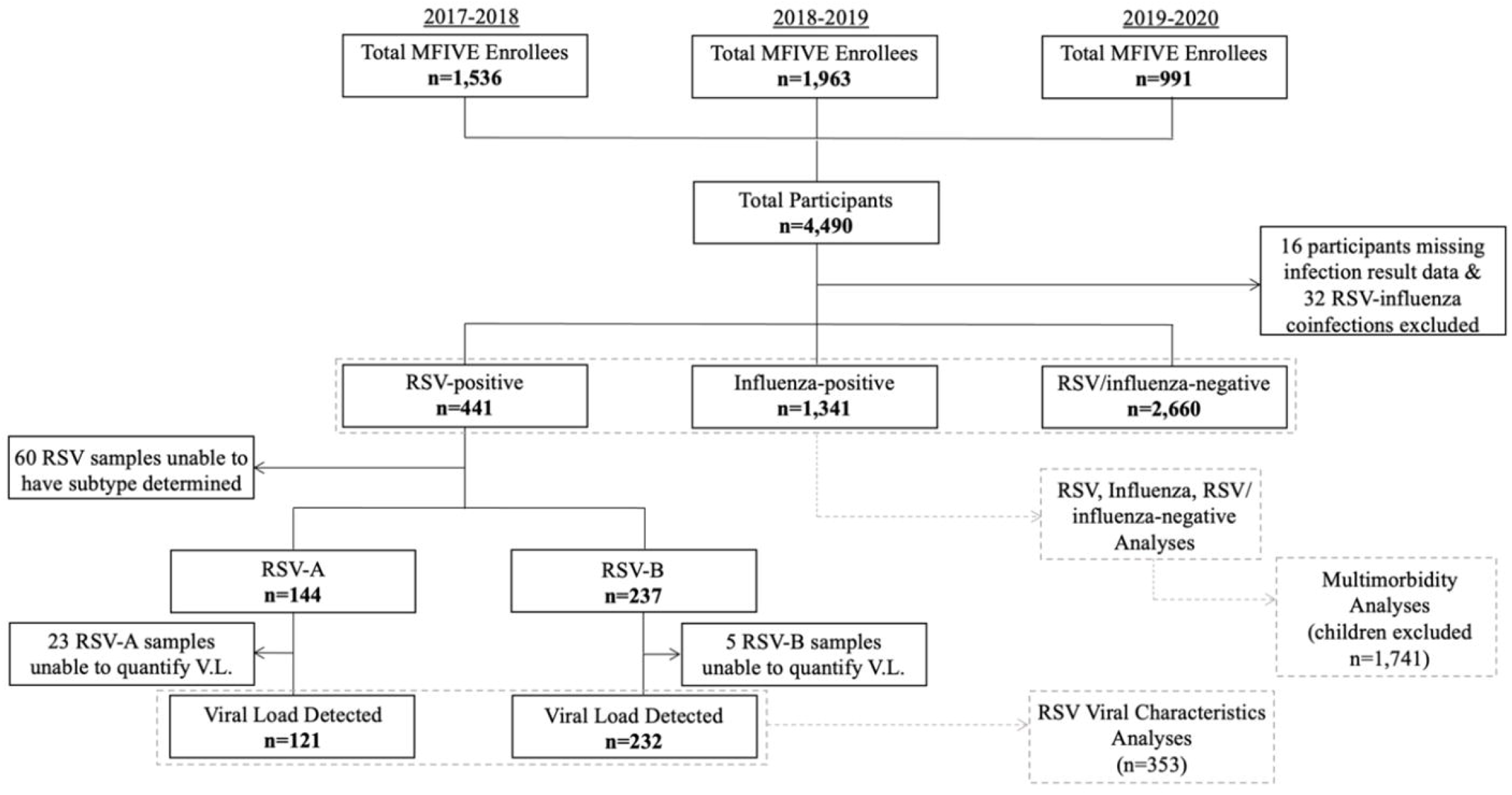
Analytic Flow Chart.

**Figure 2.**
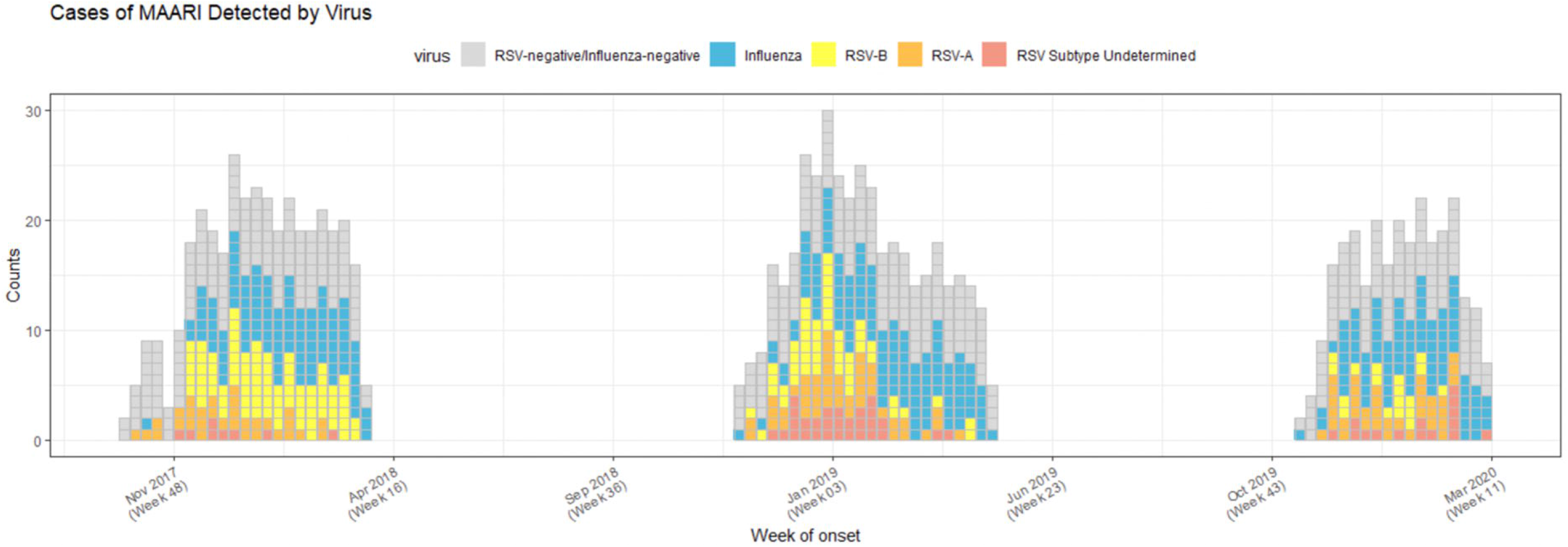
MFIVE MAARI Epidemiologic Curve by Detected Virus Type (2017-2020)

The annual proportion of RSV detection among participants under 18 years of age with MAARI across 2017/18, 2018/19, and 2019/20 was 17.0%, 11.8%, and 10.4%, respectively. Among children aged 0-4 years old across all study years (n=719), 23.8% (n=171) had RSV detected, which was slightly higher than the proportion of children in that age group with influenza detected (20.0%, n=144). The annual prevalence of symptomatic RSV among adults with MAARI across included study years was 9.4%, 6.8%, and 6.9%, respectively. These estimates were comparatively lower than the annual prevalence of symptomatic influenza among adults (36.3%, 18.5%, and 31.4%, respectively) and children (32.6%, 34.7%, and 40.0%, respectively) with MAARI in our study.

The median age of those with RSV detected was younger than those with influenza detected as well as those who were negative for both RSV and influenza (Table 1). Over two-thirds of adults with RSV detected had an MWI score above zero (68.6%, n=142). In contrast, 58% of adults with influenza detected or neither RSV nor influenza detected had an MWI score above zero. Even with a significant presence of multimorbidity across all study participants, 92.6% (n=4,114) reported being in ‘Good Health’. Among adult participants (n=2,701), those with RSV (1.2%, n=33) or influenza (7.2%, n=194) detected had children under the age of twelve residing in their household, compared to 16% (n=433) of adults negative for RSV and influenza.

**Table 1.**
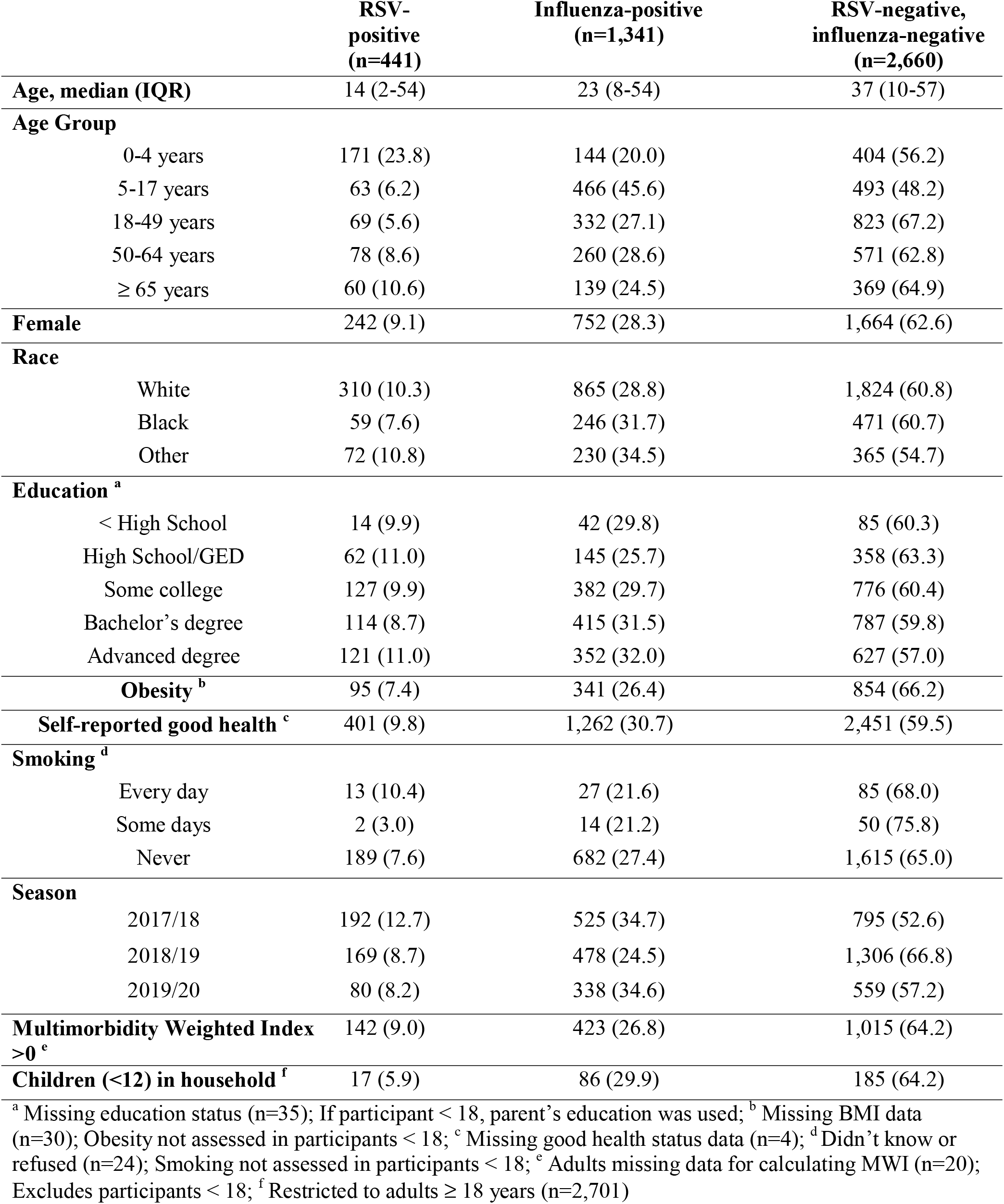
Characteristics of participants by infection detection status (2017-2020), n (row %)

|  | **RSV-**<br>**positive**<br>**(n=441)** | **Influenza-positive**<br>**(n=1,341)** | **RSV-negative,**<br>**influenza-negative**<br>**(n=2,660)** |
| --- | --- | --- | --- |
| **Age, median (IQR)** | 14 (2-54) | 23 (8-54) | 37 (10-57) |
| **Age Group** |  |  |  |
| 0-4 years | 171 (23.8) | 144 (20.0) | 404 (56.2) |
| 5-17 years | 63 (6.2) | 466 (45.6) | 493 (48.2) |
| 18-49 years | 69 (5.6) | 332 (27.1) | 823 (67.2) |
| 50-64 years | 78 (8.6) | 260 (28.6) | 571 (62.8) |
| ≥ 65 years | 60 (10.6) | 139 (24.5) | 369 (64.9) |
| **Female** | 242 (9.1) | 752 (28.3) | 1,664 (62.6) |
| **Race** |  |  |  |
| White | 310 (10.3) | 865 (28.8) | 1,824 (60.8) |
| Black | 59 (7.6) | 246 (31.7) | 471 (60.7) |
| Other | 72 (10.8) | 230 (34.5) | 365 (54.7) |
| **Education <sup>a</sup>** |  |  |  |
| < High School | 14 (9.9) | 42 (29.8) | 85 (60.3) |
| High School/GED | 62 (11.0) | 145 (25.7) | 358 (63.3) |
| Some college | 127 (9.9) | 382 (29.7) | 776 (60.4) |
| Bachelor's degree | 114 (8.7) | 415 (31.5) | 787 (59.8) |
| Advanced degree | 121 (11.0) | 352 (32.0) | 627 (57.0) |
| **Obesity <sup>b</sup>** | 95 (7.4) | 341 (26.4) | 854 (66.2) |
| **Self-reported good health <sup>c</sup>** | 401 (9.8) | 1,262 (30.7) | 2,451 (59.5) |
| **Smoking <sup>d</sup>** |  |  |  |
| Every day | 13 (10.4) | 27 (21.6) | 85 (68.0) |
| Some days | 2 (3.0) | 14 (21.2) | 50 (75.8) |
| Never | 189 (7.6) | 682 (27.4) | 1,615 (65.0) |
| **Season** |  |  |  |
| 2017/18 | 192 (12.7) | 525 (34.7) | 795 (52.6) |
| 2018/19 | 169 (8.7) | 478 (24.5) | 1,306 (66.8) |
| 2019/20 | 80 (8.2) | 338 (34.6) | 559 (57.2) |
| **Multimorbidity Weighted Index**<br>**>0 <sup>e</sup>** | 142 (9.0) | 423 (26.8) | 1,015 (64.2) |
| **Children (<12) in household <sup>f</sup>** | 17 (5.9) | 86 (29.9) | 185 (64.2) |

<sup>a</sup> Missing education status (n=35); If participant < 18, parent's education was used; <sup>b</sup> Missing BMI data (n=30); Obesity not assessed in participants < 18; <sup>c</sup> Missing good health status data (n=4); <sup>d</sup> Didn't know or refused (n=24); Smoking not assessed in participants < 18; <sup>e</sup> Adults missing data for calculating MWI (n=20); Excludes participants < 18; <sup>f</sup> Restricted to adults ≥ 18 years (n=2,701)

### Illness Outcomes

Over three-quarters of patients with influenza detected (78.5%, n=1,053) reported experiencing a fever at their enrollment visit, compared to roughly half for participants with RSV (54.2%, n=239) or neither (46.3%, n=1,231) detected. A sore throat was more often reported by those with influenza (67.3%, n=902) or neither (68.0%, n=1,808) detected compared to those with RSV detected (55.1%, n=243) (*X*^*2*^, p-value < 0.0001), and an overwhelming majority of all participants reported congestion as a symptom (87.3%, n=3,878). Nearly half of participants with influenza detected reported having all three symptoms (49.1%, n=658), and a majority of those with RSV (51.2%, n=226) or neither (46.5%, n=1,236) detected reported the presence of two symptoms (*X*^*2*^, p-value < 0.0001). Overall, 63.9% of participants completed the follow-up survey after their illness visit (n=2,838). Nearly 40% of these participants with RSV detected reported a length of illness equal to seven days or longer (Table 2). Of those who responded to the follow-up survey, 11.6% of participants with RSV, 7.7% with influenza, and 8.8% with neither detected reported seeking subsequent care (*X*^*2*^, p-value = 0.03).

**Table 2.**
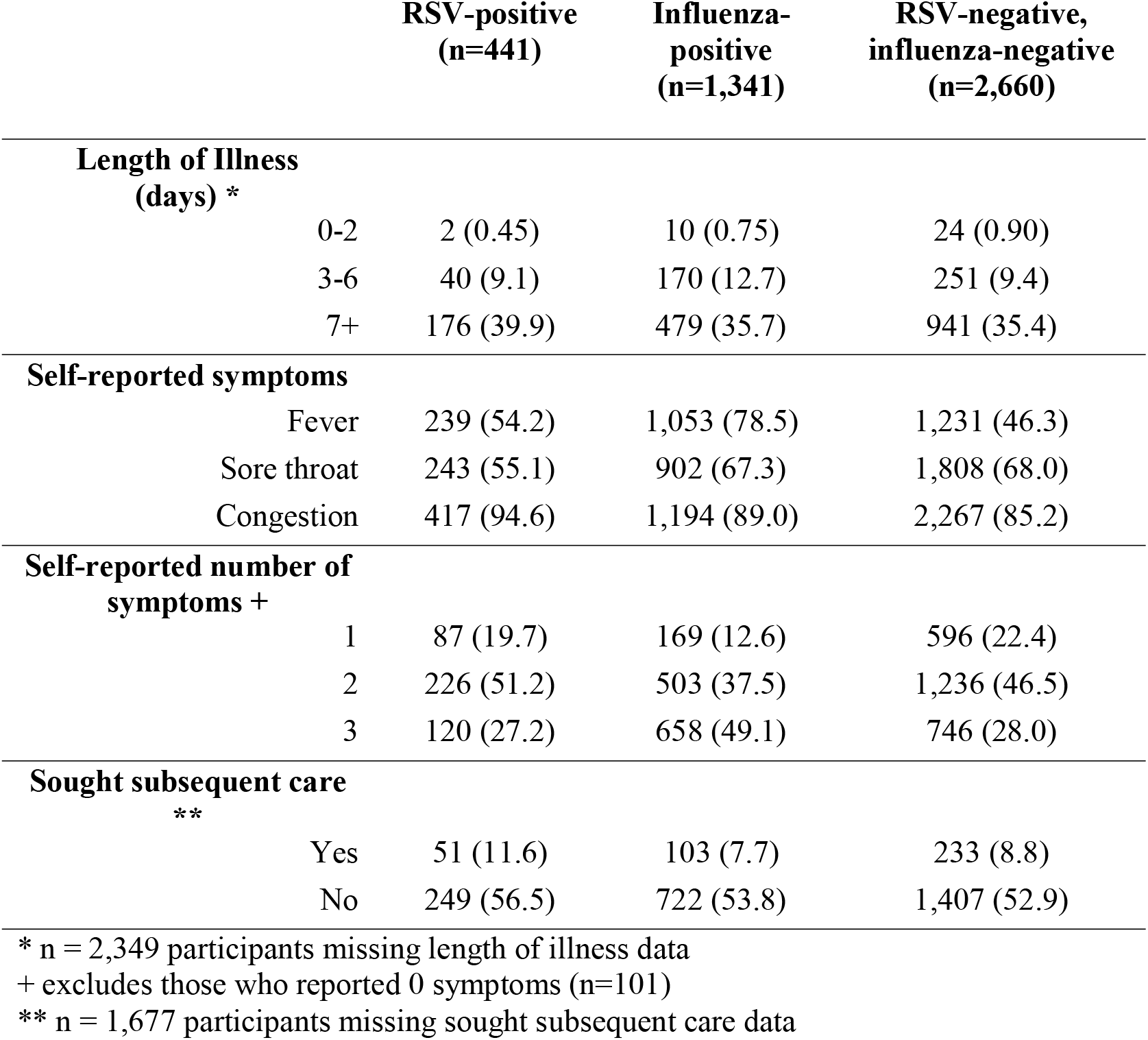
Illness characteristics by infection detection status (2017-2020), n (%)

### RSV Viral Characteristics

Among RSV-positive samples with subtyping data available (86.4%, n=381), 62.2% (n=237) were RSV-B and 37.8% (n=144) were RSV-A (Figure 1). Viral subtype was undetermined for 13.6% (n=60) of RSV specimens, and quantitative viral load was undetermined for 7.5% (n=28) of specimens and these were excluded from respective analyses. RSV-B was the most common subtype detected across these three respiratory illness seasons (Figure 2). Among samples with viral load determined (n=353), quantitated viral load ranged from 2.09×10^1^ to 1.10×10^9^ copies/mL and median viral load was 2.20×10^4^ copies/mL (log10-transformed = 4.3). Log10-transformed viral load was weakly but significantly correlated with viral subtype (*r* = 0.23, p-value < 0.0001), and RSV-B samples had significantly higher viral loads (copies/mL) when compared to RSV-A samples (Mann-Whitney test, p-value <0.0001).

After adjusting for age, gender, season, race, and time between illness onset and specimen collection, the odds were 1.88 times higher that RSV-B samples had a high viral load (defined as ≥ 2.20×10^4^ copies/mL) when compared to RSV-A samples [OR_adj_ = 1.88 (95% CI: 1.14-3.11), p-value = 0.01] (Table 3). When stratified by adults and children, the effect size became stronger in children (n=193) [OR_adj_ = 2.44 (95% CI: 1.25-4.77), p-value < 0.01], whereas it was attenuated and no longer significant in adults (n=160) [OR_adj_ = 1.22 (95% CI: 0.56-2.65), p-value = 0.61].

**Table 3.** Logistic regression analysis of viral load among those with RSV-B vs. RSV-A infection, stratified by adults and children.

|  | **Unadjusted**<br>**OR**<br>**(95% CI)** | **p-value\*** | **Adjusted**<br>**OR**<br>**(95% CI)** | **p-value\*** |
| --- | --- | --- | --- | --- |
| Overall (n=353) <sup>b</sup> |  |  |  |  |
| **Viral load**<br>**(copies/mL)** |  |  |  |  |
| ≥ 2.20x10<sup>4</sup> | 1.71<br>(1.10-2.67) | 0.02* | 1.88<br>(1.14-3.11) | 0.01* |
| 18+ (n=160) <sup>c</sup> |  |  |  |  |
| ≥ 2.20x10<sup>4</sup> | 1.26<br>(0.62-2.54) | 0.53 | 1.22<br>(0.56-2.65) | 0.61 |
| <18 (n=193) <sup>c</sup> |  |  |  |  |
| ≥ 2.20x10<sup>4</sup> | 2.48<br>(1.37-4.49) | 0.003* | 2.44<br>(1.25-4.77) | 0.009* |

<sup>a</sup> Reference group for all models < 2.20x10<sup>4</sup> copies/mL
<sup>b</sup> Adjusted for age, sex, season, race, and time between illness onset and specimen collection
<sup>c</sup> Adjusted for sex, season, race, and time between illness onset and specimen collection
\* Statistically significant at $\alpha = 0.05$

After adjusting for age, sex, race, season, viral subtype, and time between illness onset and specimen collection, those with high viral load had significantly higher odds of experiencing an extended length of illness when compared to those with a lower viral load (n=166) [OR_adj_ = 3.14 (95% CI: 1.25-7.93), p-value = 0.02] (Supplementary Table 2). After stratification by RSV subtype and adjusting for age, sex, race, season, and time between illness onset and specimen collection, participants with RSV-A detected (n=54) had a larger effect estimate [OR_adj_ = 4.88 (95% CI: 0.82-29.1), p-value = 0.08] compared to participants with RSV-B detected (n=112) [OR_adj_ = 2.30 (95% CI: 0.75-7.06), p-value = 0.15]. However, an interaction term for viral load and RSV subtype in the overall statistical model was not significant (p-value=0.61).

### Multimorbidity Analysis

The relationship between ARI and multimorbidity was examined for adult enrollees (n=2,681; Supplementary Table 1). For all three years, the most common condition categories among adults were pulmonary (20.7%), endocrine (38.4%), and cardiovascular (15.9%). The top three prevalent conditions detected across all study years were elevated cholesterol hyperlipidemia, diabetes, and asthma. Calculated non-zero MWI scores for individual RSV-positive adults ranged from 0.15-25.9, whereas scores ranged from 0.15-54.54 in influenza-positive adults and 0.15-36.57 in the negative control group. The median MWI score of RSV-positive adults in this sample higher than that in influenza-positive adults and RSV-negative / influenza-negative adults (1.62, 0.40, 0.64, respectively) (Figure 3, Supplementary Table 3).

**Figure 3.**
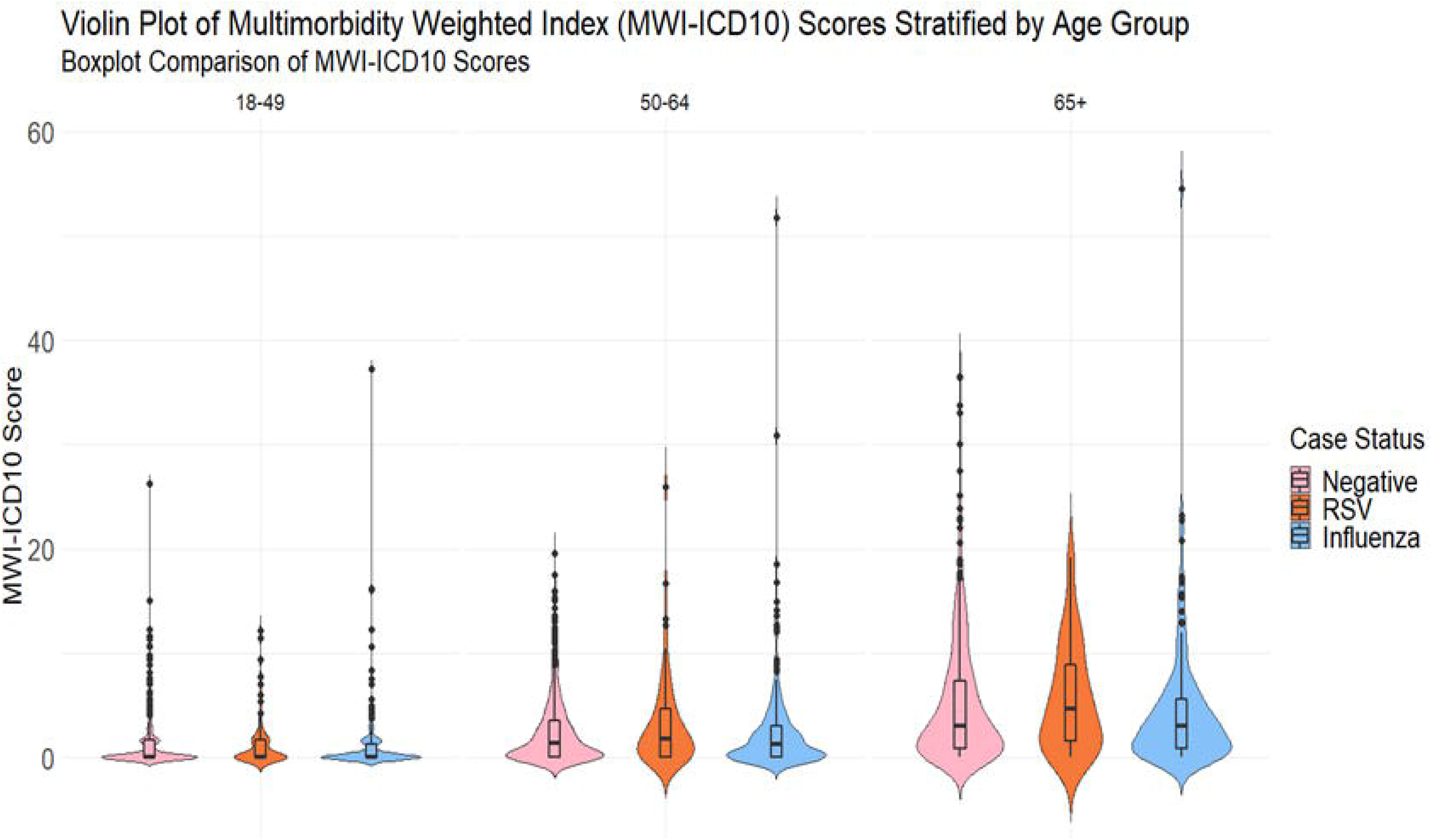
Multimorbidity Violin Plot.

Across all groups, median MWI scores were similar for those who sought subsequent care and those who did not. Adults with RSV had higher overall median MWI scores across various illness outcomes, including reporting congestion or sore throat, two symptoms total, and experiencing a length of illness more than seven days (Supplementary Table 3). After adjusting for age, sex, season, and race, there were no statistically significant associations between multimorbidity and illness outcomes for any infection group (Table 4).

**Table 4.** Adjusted odds ratios of illness outcomes among adults with multimorbidity compared to adults without multimorbidity by infection status, OR_adj_ (95% CI) and Wald p-values, (n) observations overall and included in model ^a, b^.

|  | RSV-positive | p-value | Influenza-positive | p-value | RSV/FLU Negative | p-value |
| --- | --- | --- | --- | --- | --- | --- |
| Overall <sup>c</sup> | (n=207) |  | (n=730) |  | (n=1,744) |  |
| **Extended illness** <sup>□</sup> | (n=80) |  | (n=304) |  | (n=695) |  |
| Yes | 1.63<br>(0.31-8.54) | 0.56 | 1.03<br>(0.55-1.95) | 0.93 | 0.92<br>(0.58-1.47) | 0.73 |
| **No. of reported symptoms** | (n=207) |  | (n=730) |  | (n=1744) |  |
| High ($\geq 2$) | 0.91<br>(0.43-1.93) | 0.80 | 1.36<br>(0.89-2.08) | 0.16 | 1.11<br>(0.87-1.41) | 0.40 |
| **Subsequent care** <sup>§</sup> | (n=128) |  | (n=422) |  | (n=1032) |  |
| Yes | 0.91<br>(0.29-2.86) | 0.88 | 1.07<br>(0.59-1.94) | 0.82 | 1.44<br>(0.98-2.10) | 0.06 |

<sup>a</sup> Reference group for extended illness and sought subsequent care outcomes was 'No', reference group for number of reported symptoms was 'Low ( $< 2$ )'
<sup>b</sup> Adjusted for age, sex, race, and season
<sup>c</sup> One influenza-positive adult and 19 adults negative for both did not have data to calculate an MWI score
<sup>□</sup> length of illness data missing for n=1,049 negative, n=127 RSV-positive, and n=426 influenza-positive adults.
<sup>§</sup> sought subsequent care data missing for n=712 negative, n=79 RSV-positive, and n=308 influenza-positive adults.

## Discussion

In this population presenting for outpatient care for symptomatic respiratory illness, we identified a substantial prevalence of RSV among both children (13.4%) and adult (7.7%) populations, respectively, with both RSV-A and RSV-B co-circulating through all three years of the study. Nearly one-quarter of children aged 0-4 years old (23.8%) with MAARI during the study had RSV detected. Similarly, Jackson et al. found the highest mean annual incidence of medically attended RSV among children between the ages of one and four years old [7]. We found viral load differed by subtype, and there was no difference in sample timing between RSV-A and RSV-B specimens. Viral load was also associated with reported length of illness.

Studies of hospitalized infants predating the introduction of molecular methods, have suggested that RSV-A was more likely to cause severe illness or necessitate intensive care when compared to RSV-B [24–26]. In contrast, and in line with multiple early studies, Monto and Ohmit found no differences in illness characteristics between the two subtypes in a community setting [27–31]. More recent studies describing the relationship between RSV subtype, viral load, and illness severity have focused primarily on hospitalized infant populations and report varied results. In contrast to our findings, a study of hospitalized infants by Rodriguez-Fernandez et al. found that RSV-A samples had significantly higher quantitative viral loads when compared to RSV-B samples [32]. Walsh et al. assessed RSV illness severity in infants – measured via hospitalization, ICU admission, and the need for ventilation – and concluded viral load did not differ by illness severity [33].

As age increases so does the presence of chronic conditions, putting individuals at greater risk of hospitalization or death due to RSV-associated illness. Specifically, diabetes and chronic obstructive pulmonary disease have long been recognized as risk factors for severe respiratory viral disease. The presence of these conditions has frequently been used to prioritize vaccination, particularly in adults under 65. As vaccine development for RSV progresses, it is increasingly important to recognize the presence of RSV in individuals with chronic disease, the total burden of which is represented by multimorbidity. We have previously found that adults hospitalized with RSV had significantly higher median Charlson Comorbidity Index scores (3 vs. 2, p-value < 0.001) when compared to those with influenza detected [3]. Although adults with RSV were younger than adults with influenza in our ambulatory study population, we found that adults seeking medical care for symptomatic RSV-associated illness had significantly higher median multimorbidity scores when compared to adults seeking medical care for symptomatic ARI with influenza or neither RSV nor influenza detected. This indicates that underlying multimorbidity, and the functional impact of the multimorbidity, is important to consider when identifying adults at risk for RSV-associated MAARI, and this population may benefit from priority vaccination against RSV in the future.

The inclusion of influenza-positive and RSV-negative, influenza-negative participants allowed us to make meaningful interpretations of RSV risk factors and severity. The MWI – as opposed to Charlson Comorbidity Index or Elixhauser Method – provides a more meaningful metric for assessing the impact of multimorbidity for this analysis because it accounts for the co-occurrence of multiple conditions and is validated against outcomes other than mortality, including physical functioning and cognitive status, which are more relevant to this ambulatory care population. Additional strengths of this study include prospective screening and enrollment of participants who met a pre-established MAARI case definition as well as the use of highly sensitive and specific molecular testing for virus detection and RSV subtype and viral load determination.

Our analysis underscores RSV as a substantial cause of MAARI among adults and children in an ambulatory care setting and provides additional insight into the complex relationships between viral characteristics and illness characteristics. Our findings reinforce that, similarly to influenza, consistent differentiation of RSV subtype is warranted to improve RSV surveillance and inform future vaccine development and implementation. Further, we provide data supporting multimorbidity as a significant contributor to RSV-associated MAARI among outpatient adults, with particularly notable impacts among adults under 65. Increasing efforts to regularly include older adults and those with multimorbidity in the identification and management of RSV-associated MAARI in the outpatient setting may reduce disease burden as well as subsequent care utilization.

## Data Availability

All data produced in the present study are available upon reasonable request to the authors
Supported in part by a research grant from the Investigator-Initiated Studies Program of Merck Sharp & Dohme Corp. The opinions expressed in this paper are those of the authors and do not necessarily represent those of Merck Sharp & Dohme Corp.
The data was previously presented at ReSViNET 2021.

