## Supplementary Materials for "Epidemiology of RSV-A and RSV-B in Adults and Children with Medically-Attended Acute Respiratory Illness over Three Seasons"

Supplementary Table 1 Characteristics of adults in multimorbidity analyses by infection detection status (2017-2020), n (row %)

|  | **RSV-positive**  **(n=207)** | **Influenza-positive**  **(n=730)** | **RSV-negative, influenza-negative**  **(n=1,744)** |
| --- | --- | --- | --- |
| **Age, median (IQR)** | 56 (44-68) | 52 (37-62) | 51 (37-62) |
| **Age Group** |  |  |  |
| 18-49 years | 69 (5.7) | 331 (27.3) | 811 (67.0) |
| 50-64 years | 78 (8.6) | 260 (28.7) | 567 (62.7) |
| ≥ 65 years | 60 (10.6) | 139 (24.6) | 366 (64.8) |
| **Female** | 142 (7.9) | 458 (25.4) | 1,204 (66.7) |
| **Race** |  |  |  |
| White | 146 (7.9) | 487 (26.5) | 1,206 (65.6) |
| Black | 40 (7.9) | 140 (27.5) | 329 (64.6) |
| Other | 21 (6.3) | 103 (30.9) | 209 (62.8) |
| **Education ^a^** |  |  |  |
| < High School | 4 (7.0) | 17 (29.8) | 36 (63.2) |
| High School/GED | 40 (10.2) | 92 (23.4) | 261 (66.4) |
| Some college | 64 (7.9) | 215 (26.4) | 536 (65.8) |
| Bachelor’s degree | 49 (6.6) | 211 (28.2) | 488 (65.2) |
| Advanced degree | 49 (7.6) | 192 (29.6) | 408 (62.9) |
| **Obese^b^** | 95 (7.4) | 340 (26.6) | 843 (66.0) |
| **Self-reported good health ^c^** | 175 (7.3) | 661 (27.5) | 1,565 (65.2) |
| **Smoking ^d^** |  |  |  |
| Every day | 13 (10.5) | 27 (21.8) | 84 (67.7) |
| Some days | 2 (3.1) | 13 (20.0) | 50 (76.9) |
| Never | 189 (7.7) | 682 (27.6) | 1,597 (64.7) |
| **Season** |  |  |  |
| 2017/18 | 81 (9.4) | 312 (36.3) | 466 (54.3) |
| 2018/19 | 84 (6.9) | 227 (18.7) | 904 (74.4) |
| 2019/20 | 42 (6.9) | 191 (31.5) | 374 (61.6) |
| **Multimorbidity Weighted Index >0** | 142 (9.0) | 423 (26.8) | 1,015 (64.2) |
| **Children <12 in household** | 33 (5.0) | 194 (29.7) | 427 (65.3) |
| ^†^ Adults missing multimorbidity data excluded from table (n=20)  ^a^ Missing education status (n=19)  ^b^ Missing BMI data (n=18)  ^c^ Missing good health status data (n=3)  ^d^ Smoking status, didn’t know or refused (n=24) | | | |

Supplementary Table 2 Logistic regression analysis of outcomes of interest among those with high detected viral load compared to those with low detected viral load, stratified by viral subtype, (n) observations overall and included in model

|  | **Unadjusted OR ^a^**  **(95% CI)** | **p-value** | **Adjusted**  **OR ^a, §^**  **(95% CI)** | **p-value** |
| --- | --- | --- | --- | --- |
| Overall (n=353) | | | | |
| **Extended length of illness ^b^** | (n=166) |  | (n=166) |  |
| Yes | 1.96  (0.91-4.24) | 0.09 | 3.14  (1.25-7.93) | 0.02* |
| **No. of reported symptoms** | (n=353) |  | (n=353) |  |
| High (≥ 2) | 1.39  (0.82-2.36) | 0.22 | 1.17  (0.67-2.03) | 0.59 |
| **Sought subsequent care ^c^** | (n=236) |  | (n=236) |  |
| Yes | 0.84  (0.44-1.60) | 0.59 | 1.03  (0.52-2.03) | 0.94 |
| RSV-A (n=121) | | | | |
| **Extended length of illness** | (n=54) |  | (n=54) |  |
| Yes | 4.39  (0.95-20.2) | 0.06 | 4.88  (0.82-29.1) | 0.08 |
| **No. of reported symptoms** | (n=121) |  | (n=121) |  |
| High (≥ 2) | 1.61  (0.64-4.03) | 0.31 | 1.76  (0.66-4.67) | 0.26 |
| **Sought subsequent care** | (n=76) |  | (n=76) |  |
| Yes | 0.99  (0.31-3.12) | 0.98 | 1.19  (0.31-4.53) | 0.80 |
| RSV-B (n=232) | | | | |
| **Extended length of illness** | (n=112) |  | (n=112) |  |
| Yes | 1.22  (0.47-3.20) | 0.68 | 2.30  (0.75-7.06) | 0.15 |
| **No. of reported symptoms** | (n=232) |  | (n=232) |  |
| High (≥ 2) | 1.26  (0.66-2.43) | 0.48 | 0.97  (0.48-1.95) | 0.93 |
| **Sought subsequent care** | (n=160) |  | (n=160) |  |
| Yes | 0.77  (0.35-1.68) | 0.51 | 1.09  (0.47-2.53) | 0.84 |
| ^a^ Reference group for extended illness and sought subsequent care outcomes was ‘No’; reference group for number of reported symptoms was ‘Low (< 2)’  ^§^ Adjusted for age, sex, race, season, and time between illness onset and specimen collection ^b^ n = 187 individuals missing extended illness data  ^c^ n = 117 individuals missing sought subsequent care data | | | | |

Supplementary Table 3 Multimorbidity characteristics for RSV-positive, influenza-positive, and negative-for-both adults (2017-2020)

|  | **RSV-positive (n=207)** | **Influenza-positive (n=730)** | **RSV-negative, influenza-negative (n=1,744)** | **p-value** ^a^ |
| --- | --- | --- | --- | --- |
| **MWI score** |  |  |  |  |
| Max. | 25.89 | 54.54 | 36.57 |  |
| Median (IQR) | 1.62 (0-4.9) | 0.40 (0-2.7) | 0.64 (0-3.0) | 0.0001* |
| Mean | 3.29 | 2.16 | 2.37 |  |
| **Median MWI score by reported symptom** |  |  |  |  |
| Fever | 1.33 | 0.34 | 0.63 | 0.16 |
| Sore throat | 1.53 | 0.34 | 0.34 | 0.04* |
| Congestion | 1.62 | 0.64 | 0.69 | 0.001* |
| **Median MWI score by reported number of symptoms** |  |  |  |  |
| 1 | 2.53 | 0.81 | 0.81 | 0.003* |
| 2 | 1.62 | 0.34 | 0.40 | 0.05* |
| 3 | 1.33 | 0.64 | 0.42 | 0.63 |
| **Median MWI score by length of illness (days) ^b^** |  |  |  |  |
| 0-2 | 1.62 | 0.64 | 0.81 | 0.02* |
| 3-6 | 0.15 | 0.00 | 0.34 | 0.17 |
| 7+ | 1.62 | 0.34 | 0.34 | 0.01* |
| **Median MWI score by sought subsequent care ^c^** |  |  |  |  |
| Yes | 1.62 | 1.32 | 1.30 | 0.30 |
| No | 1.62 | 0.34 | 0.34 | 0.002* |
| ^*^ p-value from Kruskal-Wallis test comparing median MWI scores  ^b^ length of illness data missing for n=127 RSV-positive, n=427 influenza-positive, and n=1,063 negative adults  ^c^ sought subsequent care data missing for n=79 RSV-positive, n=309 influenza-positive, and n=722 negative adults | | | | |
